# Aerosol transport measurements and assessment of risk from infectious aerosols: a case study of two German cash-and-carry hardware/DIY stores

**DOI:** 10.1101/2021.05.21.21257577

**Authors:** Bardia Hejazi, Oliver Schlenczek, Birte Thiede, Gholamhossein Bagheri, Eberhard Bodenschatz

## Abstract

We report experimental results on aerosol dispersion in two large German cash- and-carry hardware/DIY stores to better understand the factors contributing to disease transmission by infectious human aerosols in large indoor environments. We examined the transport of aerosols similar in size to human respiratory aerosols (0.3 µm–10 µm) in representative locations, such as high-traffic areas and restrooms. In restrooms, the observed decay of aerosol concentrations was consistent with well-mixed air exchange. In all other locations, fast decay times were measured, which were found to be independent of aerosol size (typically a few minutes). From this, we conclude that in the main retail areas, including at checkouts, rapid turbulent mixing and advection is the dominant feature in aerosol dynamics. With this, the upper bound of risk for airborne disease transmission to a susceptible is determined by direct exposure to the exhalation cloud of an infectious. For the example of the SARS-CoV-2 virus, we find when speaking without a face mask and aerosol sizes up to an exhalation (wet) diameter of 50 µm, a distance of 1.5 m to be unsafe. However, at the smallest distance between an infectious and a susceptible, while wearing typical surgical masks and for all sizes of exhaled aerosol, the upper bound of infection risk is only ∼5% and decreases further by a factor of 100 (∼0.05%) for typical FFP2 masks for a duration of 20 min. This upper bound is very conservative and we expect the actual risk for typical encounters to be much lower. The risks found here are comparable to what might be expected in calm outdoor weather.

## 1. Introduction

Humans spend most of their time indoors and it is important to keep the air in enclosed living spaces free of pollutants and other harmful substances. Airborne infectious diseases such as Severe Acute Respiratory Syndrome (SARS), avian flu and swine flu (H1N1), and more recently COVID-19 are transmitted through direct and indirect contact with an infectious person [1, 2, 3]; with the predominant route of transmission for SARS-CoV-2 being via airborne transport of respiratory aerosols [4] released from the respiratory tract of an infectious person. (In this study, we use the terms *particle(s)* and *aerosol(s)* interchangeably to refer to <100 µm particulate matter suspended in air, regardless of composition.) The size of Human aerosol emissions vary greatly in magnitude and span length scales of several decades. While moist exhaled aerosols larger than 50 µm settle by gravitational deposition on room surfaces before drying, smaller droplets dry rapidly, thereby shrink, and due to their small mass remain airborne for extended periods of time [3]. Such aerosols may contain single or multiple copies of pathogens when exhaled by an infectious, and when inhaled by a susceptible, there is a risk of infection given by the absorbed dose of pathogens [5]. In order to control and minimize airborne transmission risk by infectious aerosols, it is essential to further our understanding of aerosol dynamics and dispersion across their large spectrum of size distributions.

The spread of aerosols and the infection risk of respiratory diseases have been studied in different close contact indoor environments, such as aircraft cabins [6], buses [7], hospital wards [8, 9], and residential buildings [10, 11]. Most studies consider a well-mixed room and study fine aerosol dispersion (e.g. <10 µm) under such assumptions [e.g. see 12, 13, 14, 15, 16, 17, 18, 19, 20, 21]. In well-mixed room models and their generalizations, it is assumed that the contaminants (i.e. aerosols) are fully mixed into the whole volume of the room before they reach the destination of interest. As a result, the aerosol concentration is the same at all locations in the room and when the source is removed, the concentration decreases exponentially with time due to aerosol removing mechanisms such as deposition and air exchange [22],

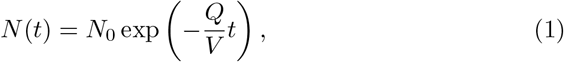

where *N*_0_ is the initial aerosol concentration, *Q* is the room clean-air supply rate typically in m^3^ h^−1^ and *V* is the room volume. The ratio *Q/V* is also known as the nominal Air Changes per hour (ACH). In cases where the mixing of the clean air with the room air is not perfect and/or when the particulate matter is removed with an imperfect filtration device, *Q* should be replaced with the effective supply rate *Q*_*E*_ = *m Q*, where 0 *< m* ≤ 1. This lead to the derivation of an effective ACH = *Q*_*E*_*/V*, which can be reformulated to calculate the *decay time τ* defined as the inverse of the effective ACH. The exponential decay shown in Eq. 1 strictly applies to an *ideal* well-mixed closed room. As we shall show later, exponential decays can also appropriately approximate the decay of aerosol concentration in open spaces, where the well-mixed assumption does not clearly apply.

There are special cases where the well-mixed assumption is a good approximation, e.g., in small spaces and when sufficient time has elapsed since the release of the pollutant. In practice, however, local emission sources always cause concentration fluctuations in the room, even if the air flow in the room is a fully developed turbulent flow. In particular, the well-mixed model fails when the volume of the room increases and/or the distance between the source and the target is large. In such situations, the aerosol concentration gradients can be taken into account with different methods, such as dividing the room into near-field and far-field regions [23, 24, 22, 25, 26, 27] or by performing detailed numerical simulations of specific scenarios [28, 29, 30].

Additionally, previous experimental work has shown that in the near-field region three different time-scales govern the aerosol concentration evolution in time, namely *t*_*α*_, *t*_*β*_, and *t*_*γ*_ [31, 32, 33]. The first time-scale, *t*_*α*_, refers to when the source is active. The duration in which the source is no longer active and the space is not yet in a well-mixed state is *t*_*β*_. And *t*_*γ*_ is the duration from when the space is well-mixed to when the aerosol concentration reaches that of the background. Turbulent mixing caused, for example, by mechanical flow devices, human presence, and thermal convection results in the space to become well mixed. If *t*_*α*_ + *t*_*β*_ ≪ *t*_*γ*_ then the well-mixed room model was proposed to be an adequate assumption for exposure risk calculations [31]. For well mixed situations, experimental studies have examined the best strategies for ventilation of indoor spaces such as schools [34], hospitals and clinics [35, 36], and homes [37]. Others studied the transport dynamics of pollutants in various confined indoor spaces [38, 39, 40] and how human presence and activity can affect these dynamics [41, 42, 43, 44]. Studies also expand upon single room confined spaces by looking at multi-zone indoor environments and how aerosols can transfer from room to room and how this transport affects aerosol concentration [45, 46, 47, 48].

To the best of our knowledge, there has not been an experimental investigation of aerosol transport in considerably large (>1000 m^3^) indoor spaces, where the well mixed assumption is known to fail. Risk of infection due to airborne disease transmission in daily-encountered environments, such as large grocery stores, shopping malls or hardware/DIY stores, have become the center of debate during the COVID-19 pandemic. The key questions in such environments are: (i) how and how quickly do local contaminants, such as respiratory aerosols produced by an infectious, disperse; (ii) what is the typical residence time of contaminants; (iii) what is the influence of store layout, presence of people, and type of ventilation; and (iv) what is the risk of infection from infectious aerosols. To answer these questions, we conducted a series of experiments in two large cash-and-carry/DIY stores in Germany. We chose these stores because they have different layouts and ventilation mechanisms. We studied the transport of aerosols similar in size to human respiratory aerosols (0.3 − 10 microns) in different locations of these stores and recorded their spatio-temporal evolution with four aerosol spectrometers. We studied high traffic locations, where close contact with people is more likely, to see how the presence of people affects aerosol transport. We also studied the effect of room layout and the possibility of aerosol transport through corridors and shelves. Our experimental observations show that exposure to local emissions, also known as near-field exposure, is the most important factor contributing to the risk of infection from airborne diseases and that the accumulation of infectious aerosols is negligible. As an example, we provide the upper bound of SARS-CoV-2 infection risk for various preventive measures such as social distancing and mask wearing. Our findings can also be applied to outdoor risk assessment in calm weather, which is similar in all respects to large indoor spaces, especially with respect to the upper bound calculations of infection risk presented here.

## 2. Methods

### 2.1. Experiments

The experiments were performed at two different hardware stores in Germany. The stores have the typical layout of similar cash-and-carry/DIY stores with high ceilings (∼10 m) and large indoor space. The two stores have different ventilation systems where Store 1 had ventilation units installed periodically on the ceiling as shown in Fig. 1(a) and Store 2 had ventilation units installed on the walls of the store, as shown in Fig. 1(b). Store 1 had a total volume of ∼190.000 m^3^ in which the garden center had a volume of ∼45 000 m^3^. The main retail area, not including the garden center, had a maximum air exchange rate of 57 800 m^3^ h^−1^ provided by the ventilation units which yield a nominal ACH of 0.4 h^−1^. Store 2 had a volume of ∼60 000 m^3^ in the main retail area and ∼40 000 m^3^ in the garden center. The measured air exchange rate was dependent on the specific location in Store 2 as proximity to ventilation units and entrances influenced the exchange rate and air currents, which made a determination of a nominal ACH not possible. Fig. 1 also shows the locations in which the aerosols were released and aerosol measurements took place. We measured specific areas of interest with higher potential for the risk of infection, such as high traffic areas with high number of customer visits and prolonged customer dwelling times. Additionally, as the store shelving may influence aerosol transport, experiments were conducted in such areas with shelf layout also shown in Fig. 1.

**Figure 1:**
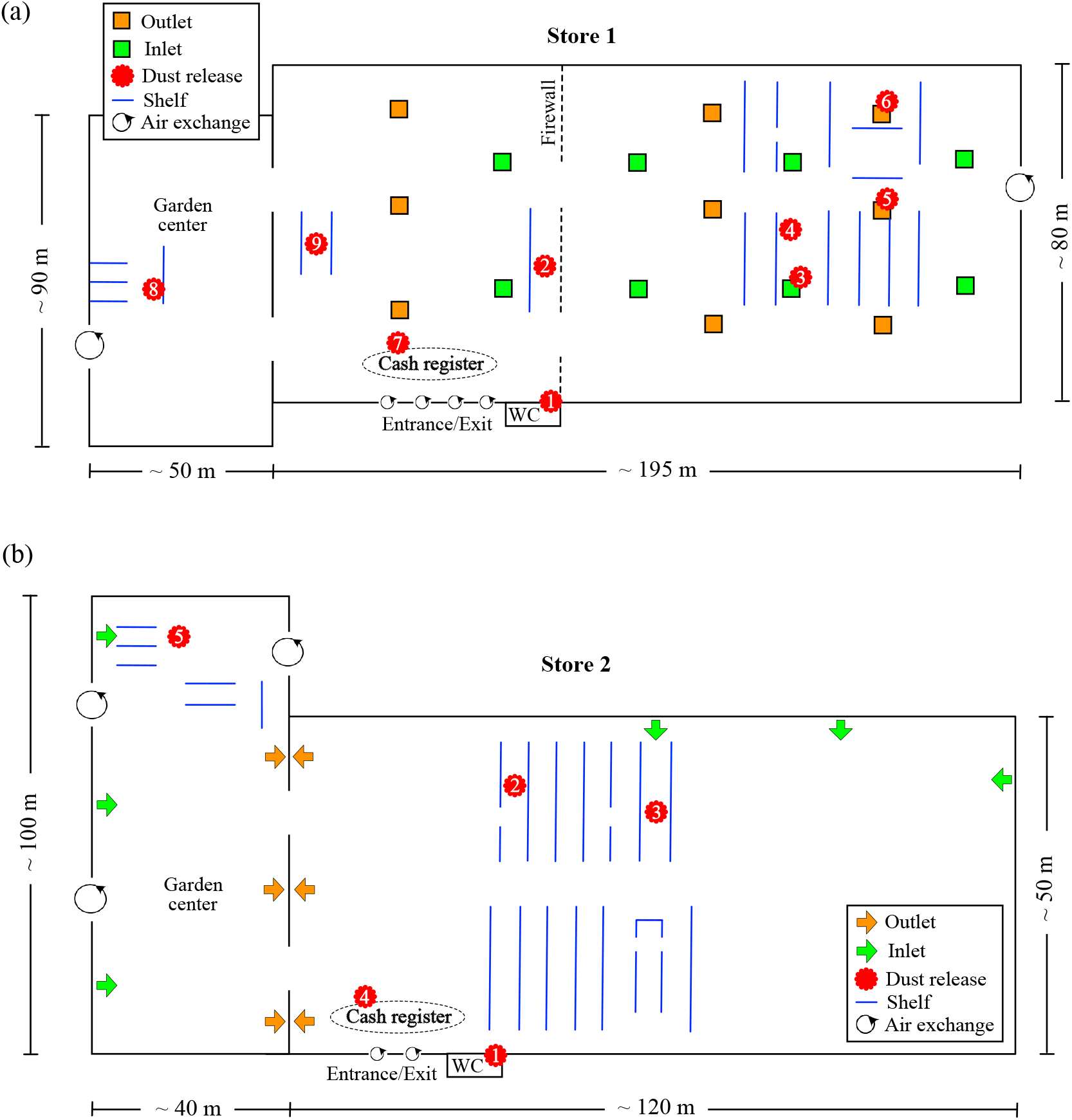
Schematics of the store layouts with ventilation units marked as well as entrances and exits that provide additional air exchange with the outside. The locations of dust release and aerosol measurement are also marked. Only the shelving around experimental cites are shown.

The aerosol measurements were conducted with four Optical Particle Sizer (OPS) spectrometers manufactured by TSI, model “Type 3330”, all of which were calibrated no later than a year before the dates on which the experiments were conducted. The individual devices are labeled as position 1, 2, 3 and 4 in each of the experiments. Each spectrometer measures particle sizes in the range of 0.3-10 µm. The devices allow for a maximum of 16 custom selected bins for size distribution analysis. In this investigation we used 16 bins that were evenly spaced on the logarithmic scale. The sampling time was set to 1 second, which yields a sample volume of 16.7 cm^3^ per sample. The OPS devices were placed on ladders or shelves during measurements at approximately the typical breathing height of an adult human (1.5-2 m above ground level). The aerosol used in the experiments was a dolomite test-dust from DMT GmbH & Co KG with diameters of less than 20 µm as specified by the manufacturer. The aerosol size distribution of this dust is relatively flat over the whole aerosol sizes and is shown in Fig. 2. The dust was released at approximately 1.0-1.5 m above ground in order to have situations similar to human aerosol generation. The release was obtained by shaking a dust-filled microfiber cloth of size 30cm x 30cm several times in a 33 L bucket and for a total duration of <5 s. Within the requirements of the experiment, this allowed for a satisfactorily controlled local release of dust, while minimizing the flow generated by the movement of the cloth itself.

**Figure 2:**
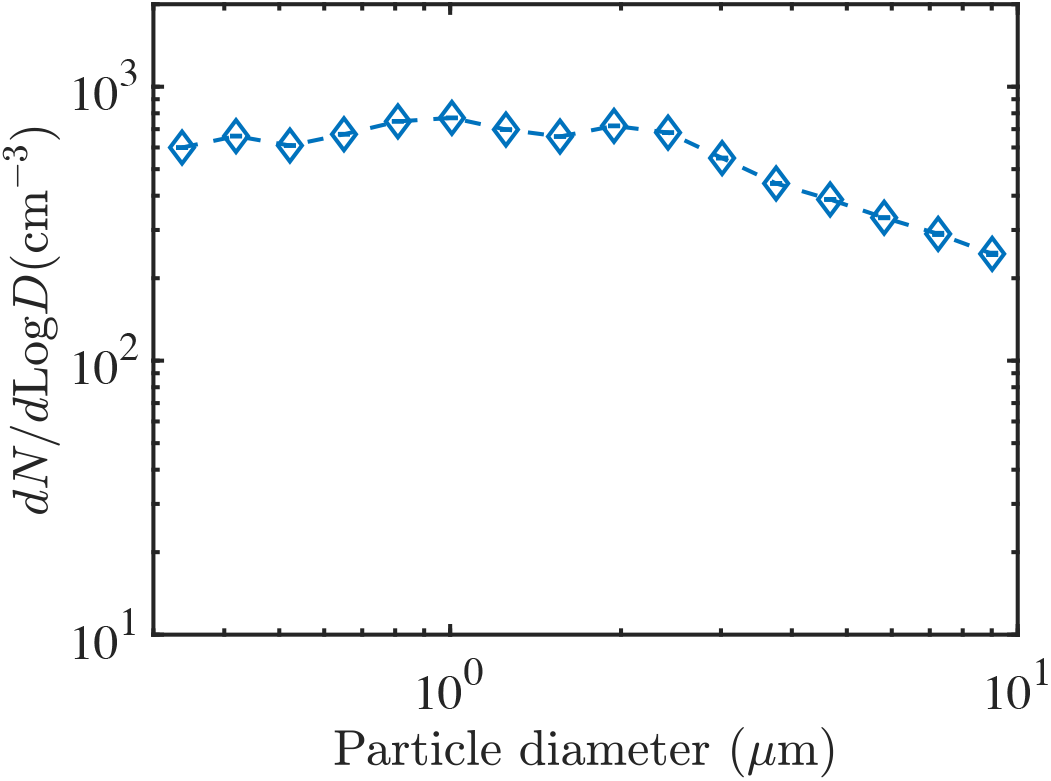
Size distribution of the DMT test dust measured immediately after the release with the OPS.

For flow visualization around inlet and outlet vents in Store 1, we used a commercial fogger (Smoke Factory GmbH), and a 5 W continuous wave (CW) frequency-doubled Nd:YAG laser that with a cylindrical lens produced a light sheet of approximately 3mm beam width and opening angel of 90°Fig. 3 shows an example of flow visualization with the laser sheet under an exhaust vent, revealing the turbulent structures in the airflow by the concentration fluctuations of the turbulently entrained fog. A thermal imager (VarioCam head from Jenoptik Laser Optik Systeme GmbH, equipped with an IR 1.0/25 LW lens) was also used to measure the temperature distribution at several experimental sites to determine whether thermally induced convective aerosol transport is possible.

**Figure 3:**
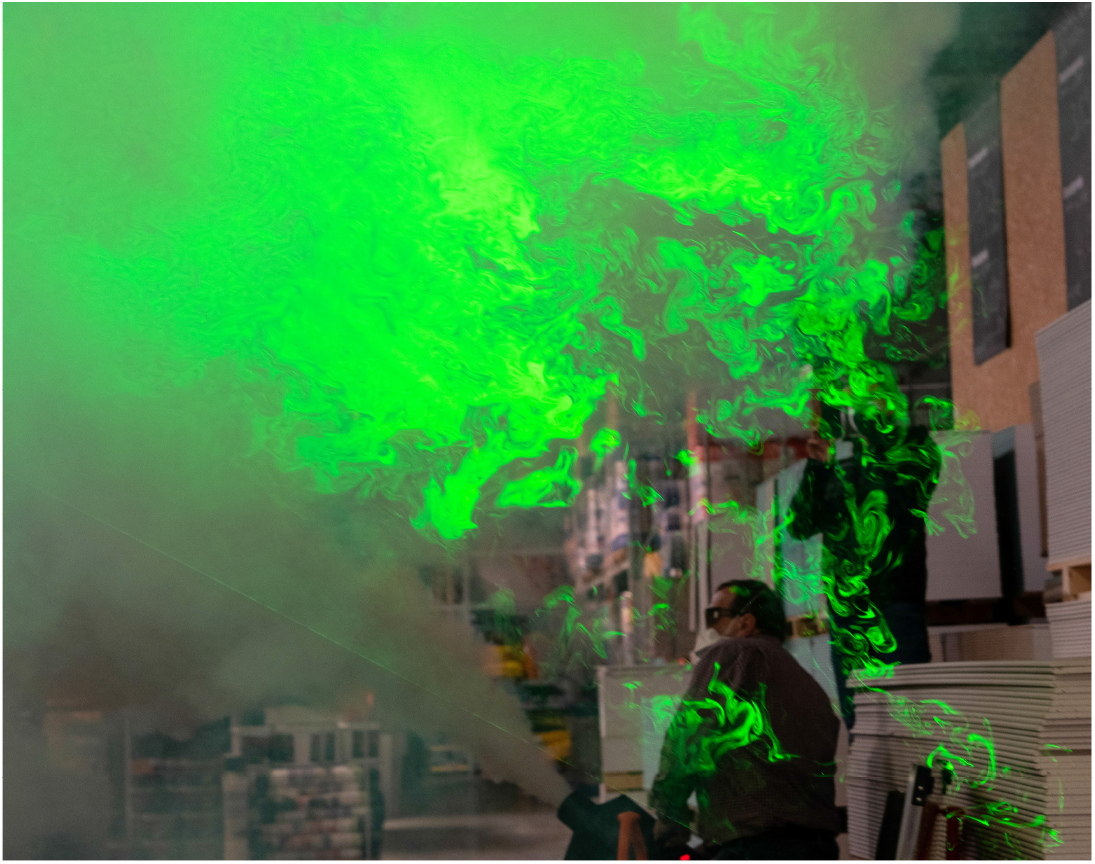
Flow visualization with a laser sheet reveals the turbulent vortices in the store air currents and their size distribution.

### 2.2. Data analysis

In this study, we examine the main features of aerosol concentration evolution using two parameters, namely the aerosol decay time and the 90% aerosol residence time. These time scales indicate how long it takes for test aerosols to become indistinguishable from background aerosols. The smaller these time scales, the faster a space returns to background conditions after aerosol release was stopped, and the lower the aerosol accumulation in space.

The aerosol decay time *τ* can only be measured if a significant portion of aerosol concentration evolution can be approximated by an exponential decay. This, however, is the case for only 42% of our measurements, i.e. 54 out of 127, which could be fitted well with an exponential similar to Eq. 1 with *r*^2^ ≥ 0.7. The starting point of such a fit was usually either the local maximum in aerosol concentration or the point in time immediately following the sharp drop that followed the maximum. The end point was either the termination of the time series or the point at which the background concentration was reached, or the point at which the concentration reached a plateau. In addition, the fit was not applied to regions where a sudden drop in aerosol concentration was observed. Some measurements required manual tuning of start and end points due to multiple peaks and other deviations from the ideal case of a simple exponential decay. We attribute these large variations to quickly changing air currents generated by the presence of customers (motion wakes and thermal convection) near the instrument.

The second parameter that parameterizes aerosol dynamics is the 90% aerosol residence time, which allows us to determine the event duration and, as shown, can be related to the aerosol decay time. The 90% aerosol residence time is calculated based on the cumulative normalized concentration evolution. By definition, the cumulative normalized concentration is a monotonically increasing function of time with values between 0 and 1. From this function, the time for reaching a certain concentration level can be determined. We use the times where the cumulative normalized concentration is 0.05 (5th percentile) and 0.95 (95th percentile) to determine the time span that covers 90% of the total aerosol concentration *t*_90%_.

It should be noted that in some experiments, the aerosol concentration at one or more measurement locations was not sufficient for quantitative analysis as the aerosol dust was carried away by weak air currents from the measurement locations. A threshold of 5% total concentration relative to the maximum total concentration was set for discrimination, and 104 of 127 data sets exceeded this threshold.

### 2.3. Infection risk

The infection risk calculations follow the methods described in [49], which were developed to estimate an upper bound for COVID-19 infection transmission risk from human aerosols in large indoor environments. Two scenarios are considered:

- **distancing scenario** in which unmasked individuals are separated by 1.5 m meters and the susceptible individual is always in the exhalation cloud of the infectious individual. Aerosol concentrations in the breathing zone of the susceptible individual are estimated to be diluted by a factor of 15 compared to concentrations generated by the infectious individual.
- **mask scenario** in which persons wearing half-face masks are in close proximity to each other, while the susceptible person inhales all exhaled air directly from the infectious person’s face mask. Only surgical and FFP2 masks are considered. Aerosol size-dependent inhalation and exhalation leakages from Bagheri *et. al*. [49] are used which are based on measurements with human subjects.

In both scenarios, the susceptible person is breathing, while the infectious person is either breathing or speaking. It should be noted that the *mask scenario* greatly overestimates the risk of infection as explained in Bagheri *et. al*. [49]. In short, the main emission from individuals wearing face masks is due to leakage flows between the mask and the face. For FFP2 masks, leakage occurs mostly at the sides of the nose and is directed upward; for surgical masks, nasal leakage and cheek leakage to the side are significant. To achieve an upper bound for infection risk all exhaled air is inhaled by a masked individual. When considering geometry alone, the upper bound given here may overestimate the risk by at least a factor of 10; when considering turbulent mixing away from the person, it may overestimate the risk by at least a factor of 100. However, because the upper bound is the most conservative and best estimate of the risk for aerosol transmission of disease, the results are presented as such.

It is assumed that both individuals are 35 years old. Gender and BMI have little effect on the estimation of the upper bound. In addition, the viral load in the respiratory fluid of the infectious individual is assumed to be constant across all aerosol sizes and equal to 1.5 ×10^8^ mL^−1^, which is close to the mean viral load for the initial SARS-CoV-2 variant but lower than those found for the B.1.1.7 variant [50]. The SARS-CoV-2 infective dose required for 63.21% chance of infection is taken to be 200. Both values are within the range reported in the literature on the current variants of the virus [49]. Aerosol size distributions for breathing and speaking activities investigated here are obtained from the HEADS database [51]. Aerosols with exhale (wet) diameter of <50 µm are considered for calculating risk of infection, assuming that larger aerosols deposit on the mask fabric or room surfaces before reaching the susceptible individual. Virus inactivation is negligible for the investigated duration of up to 60 min.

For both scenarios, it is also assumed that all aerosols inhaled by the susceptible individual have become smaller by a factor of 4 due to drying than when they were generated in the respiratory tract of the infectious individual [49]. Ventilation rates and deposition in the respiratory tract of the susceptible individual are calculated based on the ICRP lung deposition model [52], where the inhaled aerosols undergo time-dependent hygroscopic growth and under best estimates are only composed of water and NaCl. The risk of infection is calculated under the assumption of an exponential model and monopathogenic aerosols [5]. Note that for the parameters studied here, the polypathogen model yields a lower infection risk [5] and therefore the monopathogen model is best for estimating the upper bound. We also assumed that the initial absorbed dose to the susceptible individual was zero.

If the indoor space is *sufficiently large* the accumulation of virus in the background is negligible. In the case of a large open space, the threshold *sufficiently large* can be estimated such that the sum of the exhaled volume for each person for a residence time of 1 h in the room is <1% of that of the *capped room volume*. The *capped room volume* represents the estimation for an effective (virtual) room size, it is the area of the room multiplied by 2 m as the *capped height*, which is a conservative estimate of the height at which most of the exhaled cloud should dwell (in practice, this depends on the type of ventilation, the buoyancy of the exhaled cloud, and other factors). Given a typical respiratory rate of 0.5 m^3^*/*hour for breathing/speaking activities for an adult person, the occupancy density that meets our criterion is 25 m^2^*/*person.

The initial viral concentration of the air is also assumed to be zero. Note that the infection risks presented here therefore do not apply to situations in which the infectious person is in a fixed location for a long time, i.e. >1 h. In this case, infectious aerosol accumulation is possible and well-mixed zone assumptions with actual air exchange rates should be considered. For this a detailed knowledge of the individual situation is necessary. However, since the upper bound is already quite generous in estimating the risk of infection, we expect that the risk of infection in such situations will not exceed the upper bound given here for realistic situations, where some airflow and air exchange can be expected. In addition, the aerosol decay time, *τ*, should be <4 min so that the influence of viral accumulation in the space between individuals compared with that of the exhalation cloud is <10% [49]. For more details on infection risk calculations and the effects of different masks on infection risk, see Bagheri *et. al*. [49].

## 3. Results and discussion

### 3.1. Aerosol decay

Aerosol measurements were performed at different locations in the two stores to study aerosol transport in different scenarios and environments. For example, areas where customers typically take longer to find a particular item were studied, which also results in these areas having a higher number of visitors at a given time. Such locations are important for realistic calculations of infection risk in large indoor spaces, where local aerosol concentrations have the highest probability of causing infection. Measurements were also conducted in confined spaces with lower ventilation rates, such as public restrooms.

The measured decay times and 90% aerosol residence times for experiments performed in the two hardware stores are shown in Table 1. Examples of experiments with good quality exponential fits are shown in Fig. 4(a). Experiment 1 shows the aerosol concentration measured in the restroom of Store 1 where the mixing timescale is much larger than the other timescales involved (*t*_*α*_+*t*_*β*_ ≪*t*_*γ*_). Thus, the restroom can be considered a well-mixed room, where aerosol decay time was found to be long due to low ACH. In contrast, experiments 2 and 7 were conducted in the open retail areas of Store 1, where the assumption of a well-mixed space cannot be applied. Nevertheless, the decay can be well fitted by an exponential, as Eq. 1. The experiments performed in the large retail areas have much shorter decay times as compared to small closed spaces [see 49, and references therein]. As a result, in open spaces, direct exposure to the source of contaminants is more important in risk calculations than aerosols which are in the well-mixed surrounding environment. A contrasting case of an experiment with poor fits for aerosol concentration to an exponential is experiment 6, shown in fig 4(c). This section of the store was a high-traffic area and customers with shopping carts passed by during the measurement. A well-defined exponentially decaying region could not be found for this experiment and no estimate for *τ* could be obtained. However, we were still able to estimate the 90% aerosol residence time *t*_90%_ from the relative cumulative aerosol concentrations. The aerosol decay time and aerosol residence time are strongly correlated. Fig. 5 shows the relationship between *τ* and the 90% aerosol residence time for the 7.2 µm sized aerosols for experiments performed in the main retail area that have good exponential fits. The linear least-squares fits for aerosol sizes between 1 µm to 9 µm have reasonable coefficients of determination (*r*^2^ = 0.553 − 0.794) which potentially allows us to calculate an effective *τ* for cases where we have complicated flow conditions and no clear exponential decay.

**Table 1:** Aerosol decay times and 90% aerosol residence times for experiments performed in the two stores. Here S refers to the store and L to the locations identified in Fig. 1.

| Experimental values |  |  |  |  |  |
| --- | --- | --- | --- | --- | --- |
| Experiment | Location | $\tau$ range (min) | $\tau$ mean (min) | $t_{90\%}$ (min) | Description |
| 1 | S1 L1 | 13.86-86.57 | $34.37 \pm 4.94$ | $32.68 \pm 0.65$ | Restroom |
| 2 | S1 L2 | 0.82-1.99 | $1.13 \pm 0.05$ | $3.35 \pm 0.1$ | High traffic area |
| 3 | S1 L2 | 1.28-2.12 | $1.47 \pm 0.03$ | $4.13 \pm 0.16$ | " |
| 4 | S1 L2 | 3.32-15.04 | $5.82 \pm 0.31$ | $10.58 \pm 0.1$ | " |
| 5 | S1 L2 | NA | NA | $2.01 \pm 0.03$ | High traffic area, brief event |
| 6 | S1 L2 | NA | NA | $3.07 \pm 0.01$ | Out of high traffic area |
| 7 | S1 L3 | 0.40-4.86 | $1.49 \pm 0.12$ | $2.29 \pm 0.09$ | Under inlet |
| 8 | S1 L3 | 0.53-3.88 | $1.26 \pm 0.11$ | $2.6 \pm 0.11$ | Under inlet |
| 9 | S1 L4 | 3.26-5.26 | $3.78 \pm 0.14$ | $6.6 \pm 0.15$ | Between two inlets |
| 10 | S1 L4 | 1.81-4.24 | $2.49 \pm 0.07$ | $5.37 \pm 0.05$ | Between two inlets |
| 11 | S1 L5 | NA | NA | $2.37 \pm 0.06$ | Strong air currents |
| 12 | S1 L6 | NA | NA | $7.05 \pm 0.16$ | Under outlet |
| 13 | S1 L6 | 1.79-2.75 | $2.10 \pm 0.06$ | $5.3 \pm 0.08$ | Under outlet |
| 14 | S1 L7 | 0.45-1.02 | $0.55 \pm 0.04$ | $2.16 \pm 0.05$ | Cash register |
| 15 | S1 L7 | NA | NA | $2.08 \pm 0.02$ | " |
| 16 | S1 L7 | 0.59-0.80 | $0.66 \pm 0.02$ | $2.09 \pm 0.04$ | " |
| 17 | S1 L8 | 0.98-1.77 | $1.21 \pm 0.04$ | $1.84 \pm 0.07$ | Garden center |
| 18 | S1 L9 | 0.71-2.37 | $1.37 \pm 0.10$ | $4.5 \pm 0.07$ | Aisle to aisle |
| 19 | S1 L9 | 1.49-1.77 | $1.55 \pm 0.02$ | $6.14 \pm 0.05$ | Aisle to aisle |
| 20 | S2 L1 | 7.01-22.10 | $9.34 \pm 0.93$ | $14.59 \pm 0.09$ | Restroom |
| 21 | S2 L1 | 7.73-36.28 | $10.74 \pm 1.79$ | $11.68 \pm 0.09$ | Restroom |
| 22 | S2 L2 | 1.19-8.33 | $2.40 \pm 0.22$ | $4.62 \pm 0.07$ | High traffic area |
| 23 | S2 L2 | 0.71-6.25 | $1.37 \pm 0.13$ | $3.61 \pm 0.09$ | " |
| 24 | S2 L2 | 1.26-3.06 | $1.48 \pm 0.07$ | $3.69 \pm 0.06$ | " |
| 25 | S2 L2 | 1.77-8.90 | $2.46 \pm 0.26$ | $5.7 \pm 0.19$ | Aisle to aisle |
| 26 | S2 L2 | 1.96-5.61 | $2.67 \pm 0.22$ | $5.97 \pm 0.07$ | Across main aisle |
| 27 | S2 L3 | 0.55-3.59 | $0.83 \pm 0.07$ | $1.65 \pm 0.07$ | Downstream of inlet |
| 28 | S2 L3 | 0.59-2.80 | $1.12 \pm 0.11$ | $2.4 \pm 0.07$ | Downstream of inlet |
| 29 | S2 L3 | 0.48-1.07 | $0.58 \pm 0.04$ | $5.39 \pm 0.03$ | Downstream of inlet |
| 30 | S2 L4 | NA | NA | $1.72 \pm 0.06$ | Cash register, brief event |
| 31 | S2 L4 | 0.87-5.01 | $1.96 \pm 0.19$ | $3.1 \pm 0.04$ | Cash register |
| 32 | S2 L5 | NA | NA | $0.5 \pm 0.13$ | Garden center, brief event |
| 33 | S2 L5 | 0.45-1.80 | $0.59 \pm 0.09$ | $1.52 \pm 0.06$ | Garden center |
| 34 | S2 L5 | 0.99-4.41 | $1.48 \pm 0.14$ | $5.11 \pm 0.03$ | Garden center |

**Figure 4:**
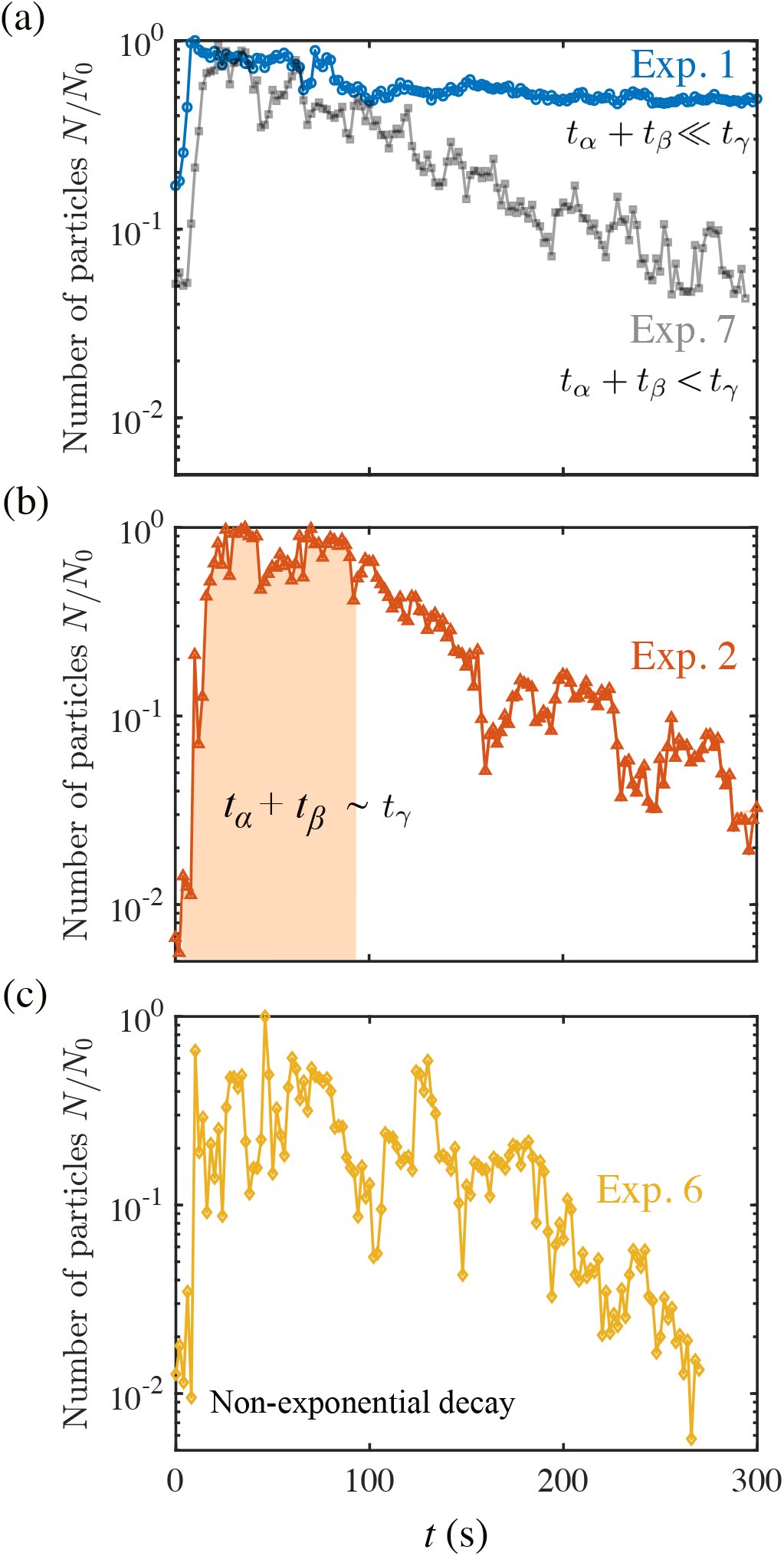
Normalized aerosol concentration for 0.90-1.12 µm sized aerosols as measured by one OPS. (a) Experiments 1 and 7 of Table 1 with an exponential decay showing different dynamics. (b) Experiment 2 where *t*_*α*_ + *t*_*β*_ ∼ *t*_*γ*_. (c) Experiment 6 where no good estimates for *τ* are found but aerosol residence time can be measured.

**Figure 5:**
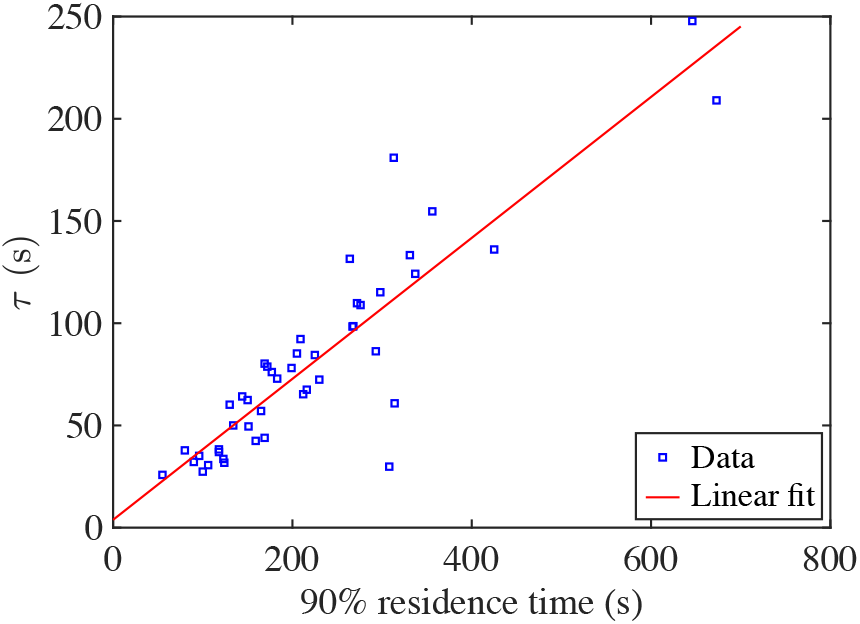
Exponential decay constant *τ* as a function of the 90% aerosol residence time for the 7.2 µm sized aerosols. The data represents 45 out of the 127 available data-sets from both stores excluding the restroom experiments that have longer *τ*.

The decay times for the different measurement locations as a function of aerosol size are shown in Figure 6(a). Aerosols remain longer air-bound in the smaller restroom areas than in the main retail areas. It was found that the restrooms can be considered well-mixed environments where aerosol dynamics are dominated by settling and deposition. In the main retail areas with open spaces and improved ventilation, the measured decay times were not strongly dependent on aerosol size and were typically 1-2 min with little variation between sites. The ventilation in Store 2 had unfiltered air intakes from outside which created a higher background concentration of aerosols smaller than 0.5 µm in diameter and resulted in longer decay times for smaller aerosols. Fig. (b) shows the PDF of measured decay times for experiments conducted in the main retail area, with most sites having a decay time of approximately 1 min. The constancy of the decay times in the main retail areas shows that, unlike in the restrooms, the aerosol dynamics in the open areas are not dominated by mixing and deposition, but by local transport through weak air currents. In our flow visualization experiments with the laser sheet, we observed that the aerosol cloud experienced strong advection-diffusion.

**Figure 6:**
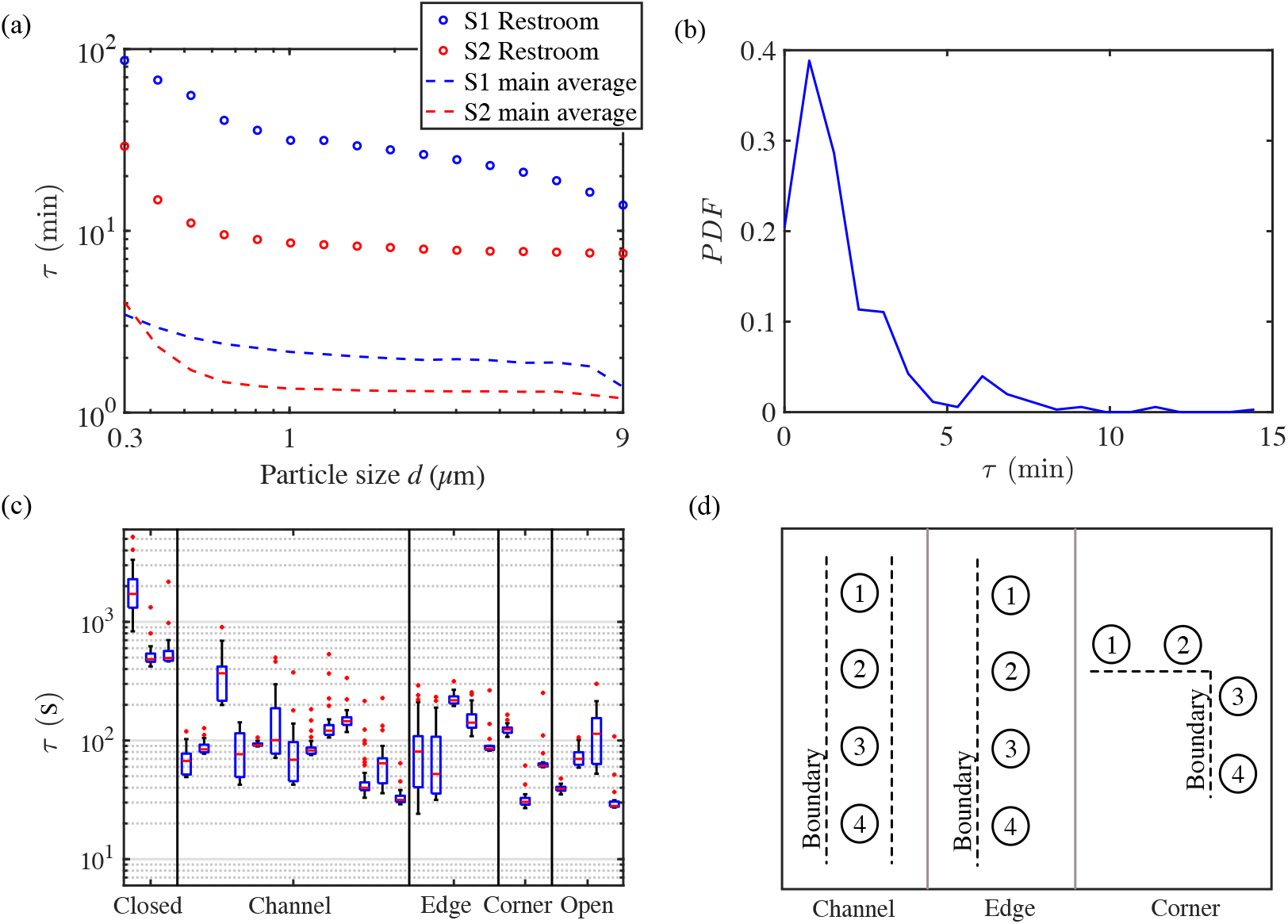
(a) Aerosol decay times *τ* as a function of aerosol size at different locations. In S2 the ventilation intake air from outside was not filtered, leading to larger decay times for small aerosols. (b) PDF of *τ* for experiments performed in the main retail area of the two stores (excluding the restrooms). (c) Box and whisker plots of *τ* for S1 and S2 where the data is grouped by the experiment location type. (d) Schematic of experiment location types.

The aerosol decay times are grouped by experiment location type and are shown in the box and whisker plots of Fig. 6(c). Fig. 6(d) shows a schematic of location types in the main retail area that have boundaries in the form of walls or shelving. *Channel* describes aisles that have two parallel boundaries, mostly shelves, on both sides. *Edge* has one boundary and the other directions are free, and *corner* describes situations where there is one parallel boundary and another boundary perpendicular to it. Open areas have no boundaries near the measurement location, such as checkout areas or garden center locations. Enclosed spaces such as restrooms showed the longest aerosol decay time of all locations studied, while the main retail spaces all showed similar and much shorter decay, with no significant difference between location types.

### High traffic locations

The results of experiment 4 are shown in Fig. 7, this location is a high traffic area with higher than average dwelling time for customers (screw section). Fig. 7(a) shows the aerosol concentration normalized by the maximum value observed in the particular experiment for aerosol sizes of 0.90-1.12 µm. The measurement devices were evenly spaced in the aisle and the aerosol dust was generated near position 1. The existence of an air current in the direction from device 1 to device 4 caused the aerosol cloud to be advected to the other measurement devices in a short amount of time. The majority of experiments performed in the open store areas were of this nature, where the aerosol dynamics was dominated by advection and diffusion due to turbulent mixing. After the release of the aerosols, the sharp rise seen in the concentrations corresponds to the arrival of the aerosols at the spectrometer locations as seen in Fig. 7(a). The decay is similar for all 4 spectrometers reflecting that the cloud is uniformly mixed in space and exponentially decays in time. Additionally, there is no significant contribution to aerosol transport from thermal convection as demonstrated by a thermal image of the experiment aisle shown in Fig. 7(b). The temperature profile of the store is relatively constant with no gradients between the floor, ceiling, and walls that would cause convective motion of air and aerosols.

**Figure 7:**
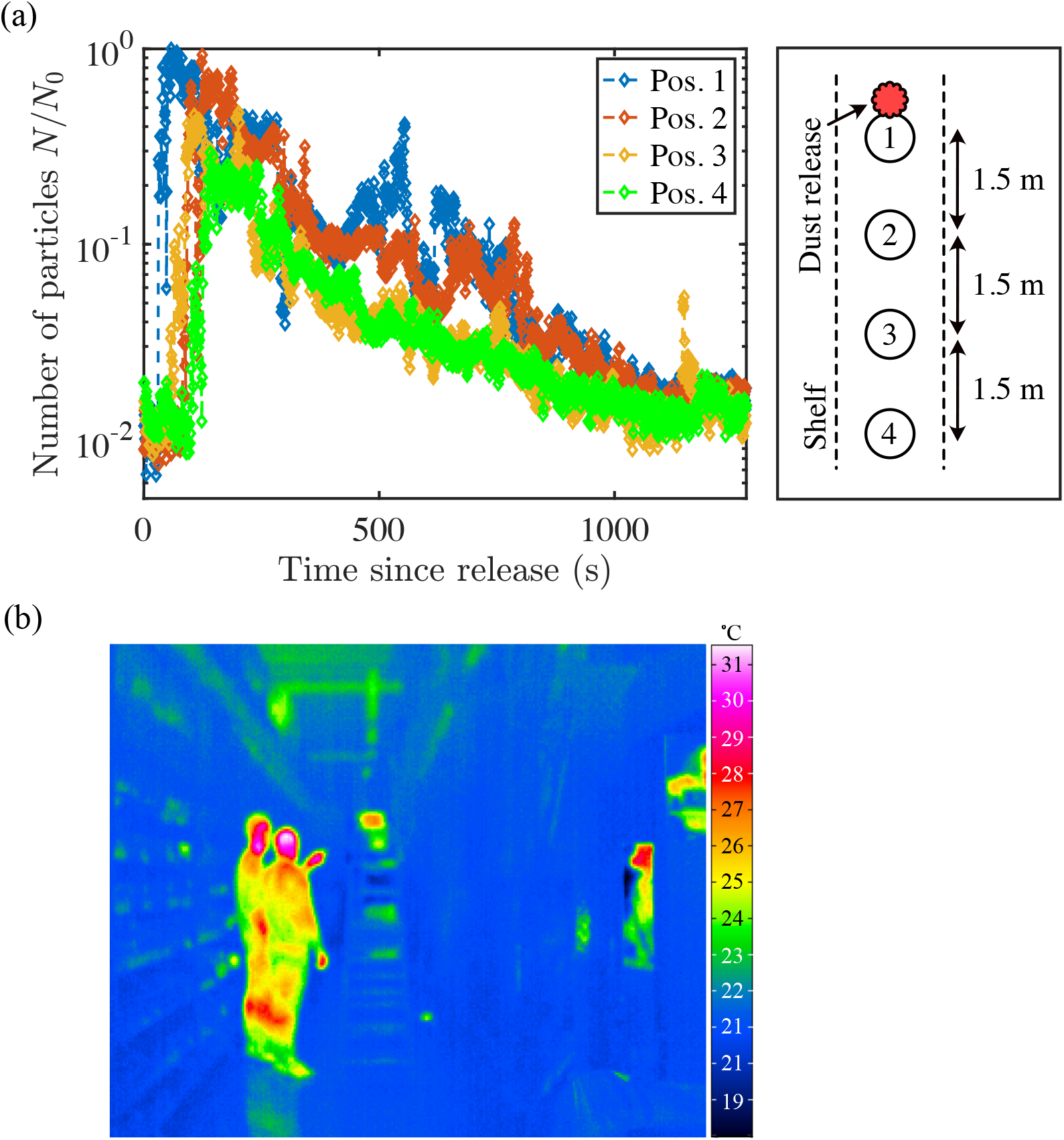
(a) Normalized aerosol concentration for aerosols of size 0.90-1.12 µm at different positions in the aisle for experiment 4. (b) Thermal camera image of an aisle with two people standing in the aisle. The temperatures in the store are relatively constant with no significant contributions to aerosol transport from thermal convection.

We have also examined high traffic, long customer dwelling time areas for cases with and without people present in the aisle. The experiments with these conditions were performed in Location 2 of Store 2 as shown in Fig. 1(b). Fig. 9(a) shows the case without people present in the aisle where the measured aerosol concentrations have been normalized by the observed maximum for aerosol sizes of 0.90-1.12 µm. For the case without people present, we found similar results to those of Location 2 in Store 1. Fig. 9(b) shows the same location but with 5-6 people present in the aisle when the aerosol dust was generated. The presence of people changes aerosol transport in a significant way, where depending on the location of individuals, a person may obstruct the aerosol transport path in the aisle. The movement of people can also create wakes and cause re-suspension and vortical transport. This demonstrates the locality of aerosol dynamics and how the aerosol does not become well-mixed in large environments. We can see this effect in the measurement made at Pos. 4 of Fig. 9(b) where the aerosol reaches Pos. 4 at a much later time and the concentration only slightly goes above the background. Furthermore, human presence can cause the incoming aerosol to be carried upward via convective flow. The displaced volume of air by the convective boundary layer of a human is *πr*^2^*v* = 0.025 m^3^ s^−1^, which is calculated by multiplying the area of a human with radius *r* = 0.2 m with the typical human convective boundary layer flow velocity of *v* = 0.2 m s^−1^ [41].

### Cross-aisle aerosol transport

Aerosol transport from aisle to aisle is also of interest. To study the paths and routes that aerosols take to move from one aisle to the next, we conducted experiments at Location 9 of Store 1. From Fig. 8(a) we see that the aerosol is only slightly transported to the neighboring aisle but mostly remains local to the area in which it was released. Here we have two spectrometers in the aisle where the aerosol dust was generated, and two spectrometers in the adjacent aisle showing aerosol transport to the next aisle. Fig. 8(b) shows the results for where we had one sepectrometer in the aisle where the aerosol is generated and 3 spectrometers in the neighboring aisle stacked on top of each other. Fig. 8(b) shows that most aerosols are detected at approximately the same height as where the dust was generated. Thus, the aerosol did not travel over or around the shelving but rather through perforations in the shelf itself. Similar to the case where people were present in the aisle, the shelving acts as a barrier to aerosol transport. The aerosol remains local to the location of release and does not become strongly well-mixed and spread to neighboring areas.

**Figure 8:**
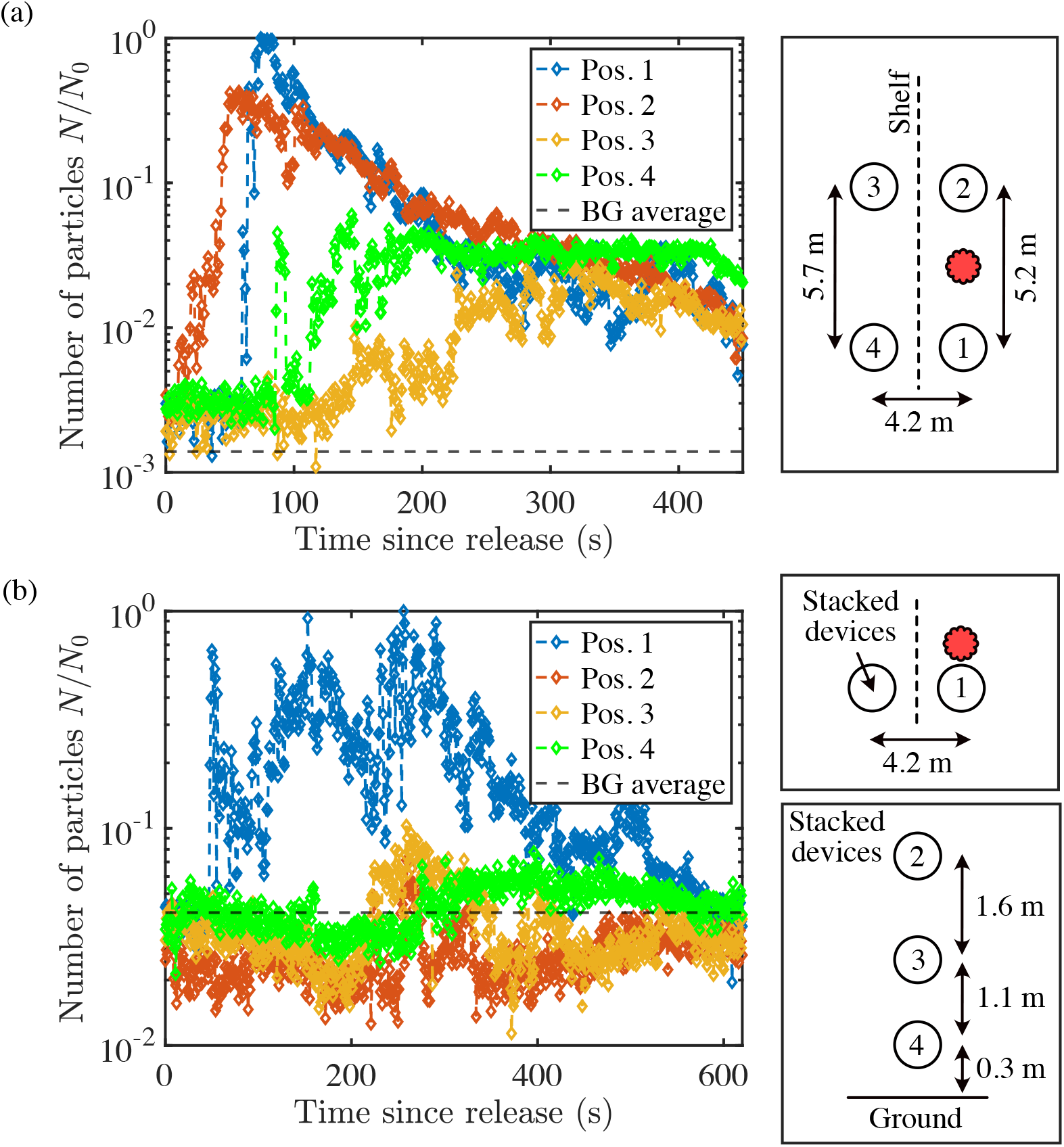
Aerosol dispersion from aisle to aisle for aerosols of size 0.90-1.12 µm. (a) Initial experiments with all measurement devices at the same height. (b) Experiment with measurement devices at various heights in the neighboring aisle to where the aerosol dust is generated as a way to determine the aerosol transport path. The concentrations have been normalized by the maximum measured concentration in the particular experiment.

### 3.2. Infection risk for SARS-CoV-2

In summary, the decay time *τ* is short in large indoor environments and is much longer for poorly ventilated confined spaces. The aerosol decay time is inversely related to the volume in which the aerosol is released and the air change per hour (ACH) of the indoor space. As the room size and ACH increase, the decay rate of aerosols decreases which is also observed by Mage and Ott [31] where they report *τ* ≈ 2.6min in the large indoor space of a tavern (*V* ≈ 550 m^3^). The short decay times measured in large indoor spaces suggest that existing well-mixed aerosols in the environment do not play a critical role in infection risk calculations and that the strongest probability of infection is caused by direct contact with an infectious individual and their respiratory droplet emissions.

The COVID-19 infection risk as a function of time is shown in Fig. 10 for distancing and mask scenarios. For each activity of the infectious individual, the highest risk of infection occurs for the distancing scenario. Being in the exhalation cloud of an unmasked speaking infectious individual for only a few minutes even at a distance of 1.5 m is enough to pose a >90% risk of infection to the susceptible individual. In all other cases, the absorbed viral dose is small enough such that the risk of infection follows a linear trend in time. Interestingly, risk of infection for speaking mask-SS is very similar to breathing-distancing-1.5m, both with a slope of 0.23% per minute reaching ∼14% after 60 min. Breathing mask-SS is associated with 3.6% of infection risk after 60 min, i.e. a slope of 0.06% per minute. When FFP2 masks are used, mean infection risk is <0.2% after 60 min of exposure regardless of the infectious individual activity.

**Figure 9:**
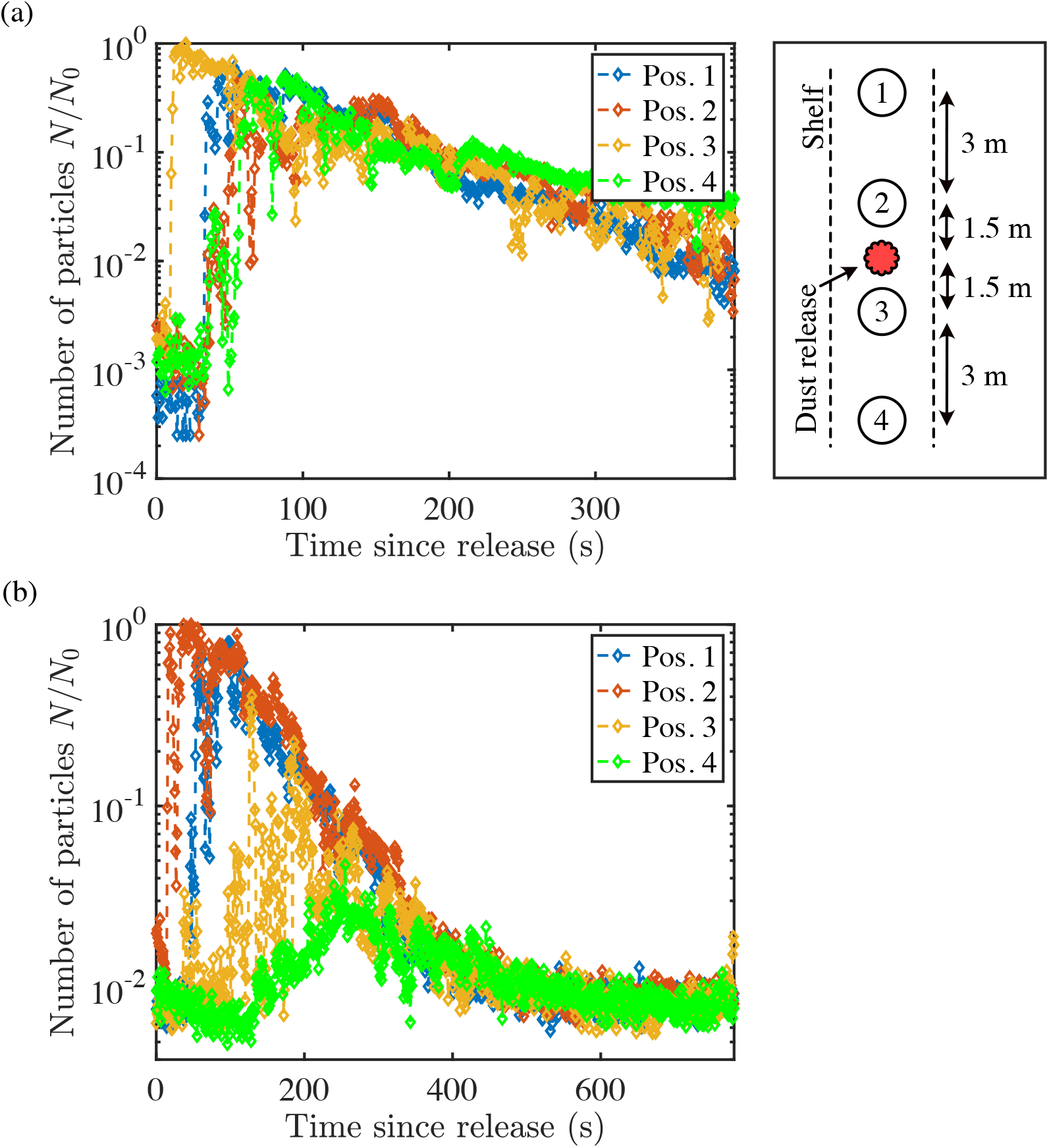
Aerosol dispersion in an aisle for aerosols of size 0.90-1.12 µm (a) with and (b) without people. The concentrations have been normalized by the maximum measured concentration in the particular experiment.

**Figure 10:**
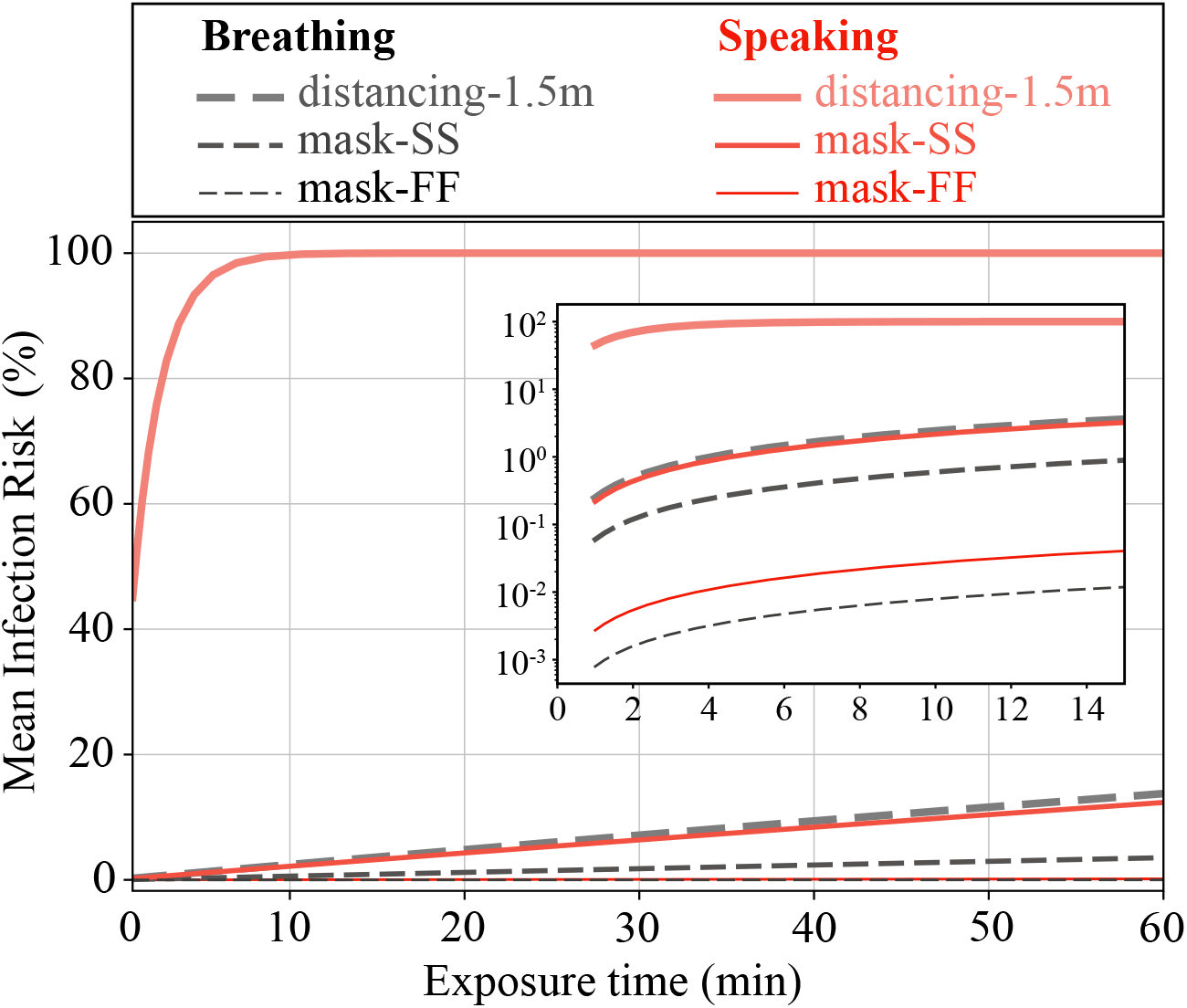
Mean infection risk as a function of time for when both infectious and susceptible individuals are breathing and for when an infectious individual is speaking to a breathing-only susceptible individual. We consider the cases where there is a 1.5 m distance between the two unmasked individuals and when different face half-masks are used with no distancing, see section 2.3 for more details on the models and assumptions. Mask-SS is the scenario in which both infectious and susceptible individuals are wearing typical surgical masks, while mask-FF represents the case in which both wear typical FFP2 masks. The x-y axes in the inset plot show the same quantities as those of the main plot but with a zoomed-in x-axis (1-15 min) and a y-axis in logarithmic scale.

If we consider 1% risk of infection as the upper threshold for a scenario to be considered *low risk*, various mitigation strategies can be implemented for typical social interactions that include normal speaking and breathing within large indoor spaces. The safest strategy between the investigated scenarios is to use reliable FFP2 masks for both infectious and susceptible individuals. This is associated with a risk of infection that is well below the low-risk threshold, even for prolonged interactions. If only (typical) surgical masks are worn by individuals, the duration of exposure should be shortened as much as possible. According to our conservative estimates, the duration at which the risk of infection reaches 1%, in this case, is approximately 4 min. Nonetheless, the safe exposure duration for mask scenarios can be increased by a factor of 10 or even more, as we greatly overestimate the risk of infection for mask scenarios. This is justified considering that when a face mask is worn, most of the leakage goes up and away from the subjects through the nosepiece. In addition, exhaled air passes through a large area of the mask fabric with a significant reduction in flow velocity, which leads to large dilution even at short distances. If masking is not possible, speaking is an extremely high risk activity even at a distance of 1.5 m, while nonvocal activities such as passive breathing are safe only up to a duration of 4 min.

These results indicate that masking is by far more efficient in reducing infection risk of transmission compared to social distancing in large indoor environments as shown also by [49]. For example, being in the exhalation cloud of a speaking infectious person for 4 minutes at a distance of 1.5 m is associated with an infection risk 90 times more than when both individuals have surgical masks and are directly next to each other. For breathing activities, the differences between mask-SS and distancing scenarios is four-fold, which is comparatively not as prominent but still significant. While with FFP2 masks the risk decreases by a factor of 100 compared to similar distancing scenarios. Overall it can be concluded that in order to reduce risk of infection between individuals in large indoor environments, masking is the only safe option. This conclusion also applies to outdoor spaces, which is the limiting case of large indoor environments [49].

## 4. Conclusions

We measured aerosol transport and aerosol dynamics in two German cash- and-carry hardware/ DIY stores to better understand the factors that are important for disease transmission via human respiratory aerosols in large indoor spaces. The aerosol used in our experiments had a similar size distribution to human respiratory aerosols, ranging from 0.3 µm to 10 µm. We took measurements in various locations that pose a high risk of infection from airborne diseases, such as high-traffic areas and restrooms. To determine the appropriate method for calculating the infection risk of infectious aerosols distributed in an environment, we measured the decay and residence time of the aerosol cloud for our experiments.

Small enclosed spaces with poor ventilation, such as restrooms, had long aerosol decay times, i.e. the aerosols remained airborne for a long time. Such environments can be considered well-mixed, with the history, i.e. the temporal accumulation of infectious aerosols in the room, governing the risk of infection. In the much larger retail areas of the two stores, the decay and residence times were much shorter. The decay times calculated in the large retail areas were found to be independent of the aerosol size and the location in the store, such as shelved areas, at the cash registers, or in open garden areas. This independence shows that aerosol dynamics are dominated by rapid turbulent mixing and advection.

We conclude that the large, well-ventilated areas with rapid aerosol decay times are not fully mixed and the leading factor determining the upper bound for the risk of disease transmission is direct exposure to aerosol emissions from an infectious. This assumption applies in the case where the background environment is “free” of infectious aerosols, which means that the occupancy must not be too large. To get an upper bound on this, we know that a person typically exhales 0.5 m^3^ of air per hour. If the person is infectious, then for the contribution to be insignificant in terms of aerosol accumulation in space, we estimate that it must be <1% of a volume that is capped to the height of a person, i.e. 2 m. This then leads to a required 25 m^2^/person.

Finally we report the upper bound of disease transmission of COVID-19 for scenarios of social distancing and wearing half-face masks. For exhaled (wet) aerosol sizes of up to 50 µm from speaking, we find that a distance of 1.5 m between unmasked individuals is unsafe with a 90% risk of infection after 4 minutes of exposure. However, even at the smallest distance between two individuals wearing surgical masks, the maximum infection risk is ∼5% after 20 minutes of exposure which decreases further by two orders of magnitude when masks are switched to FFP2 type masks.

The calculated upper bounds for infection risks are intentionally chosen to represent the most conservative estimate. In realistic situations, we expect the infection risks reported here to be significantly lower for typical encounters, i.e., to be lower by at least a factor of 10 to 100. The results presented here can also be applied to outdoor scenarios with calm weather, which are similar in all respects to a large, well-ventilated indoor environment. Finally, we note that the infection risks calculated in this study only consider direct exposure via inhalation of exhaled infectious aerosols and we do not consider the risk of indirect exposure and contact with fomites.

## Data Availability

available on request for at least 10 years after publication

## Acknowledgments

We thank the Handelsverband Heimwerken, Bauen und Garten (BHB) for bringing this issue to our attention and for providing financial support to part of this work. The open and unrestricted access to the hardware stores, the communication with the employees, as well as the unrestricted access to information and support have not only made our research project possible, but have also made it much more simple. We thank BMBF for funding within the project B-FAST (Bundesweites Netzwerk Angewandte Surveillance und Teststrategie) (01KX2021) within the NUM (Netzwerk Universitätsmedizin) and the Max-Planck-Gesellschaft. The sponsors had no influence on the study design, data collection and analysis, decision to publish, or preparation of the manuscript.

The authors would also like to thank Alexei Krekhov, Yong Wang, and Taraprasad Bhowmick for insightful discussions regarding solutions to the three-dimensional complex source advection-diffusion problem.

## References

[1] R. Zhang, Y. Li, A. L. Zhang, Y. Wang, M. J. Molina, Identifying airborne transmission as the dominant route for the spread of COVID-19, Proceedings of the National Academy of Sciences 117 (26) (2020) 14857–14863.

[2] WHO Team, Transmission of SARS-CoV-2: implications for infection prevention precautions, WHO Scientific Brief (2020).

[3] M. L. Pöhlker, O. O. Krüger, J.-D. Förster, T. Berkemeier, W. Elbert, J. Fröhlich-Nowoisky, U. Pöschl, C. Pöhlker, G. Bagheri, E. Bodenschatz, J. A. Huffman, S. Scheithauer, E. Mikhailov, Respiratory aerosols and droplets in the transmission of infectious diseases (2021). 2103.01188.

[4] N. Leung, Transmissibility and transmission of respiratory viruses, Nature Reviews Microbiology (2021).

[5] F. Nordsiek, E. Bodenschatz, G. Bagheri, Risk assessment for airborne disease transmission by poly-pathogen aerosols (2021). 2011.14118.

[6] M. P. Wan, G. N. S. To, C. Y. H. Chao, L. Fang, A. Melikov, Modeling the Fate of Expiratory Aerosols and the Associated Infection Risk in an Aircraft Cabin Environment, Aerosol Science and Technology 43 (4) (2009) 322–343.

[7] S. Zhu, J. Srebric, J. D. Spengler, P. Demokritou, An advanced numerical model for the assessment of airborne transmission of influenza in bus microenvironments, Building and Environment 47 (2012). 67–75, international Workshop on Ventilation, Comfort, and Health in Transport Vehicles.

[8] J. Ren, Y. Wang, Q. Liu, Y. Liu, Numerical Study of Three Ventilation Strategies in a prefabricated COVID-19 inpatient ward, Building and Environment 188 (2021) 107467.

[9] H. Qian, Y. Li, Removal of exhaled particles by ventilation and deposition in a multibed airborne infection isolation room, Indoor Air 20 (4) (2010) 284–297.

[10] N. Gao, J. Niu, M. Perino, P. Heiselberg, The airborne transmission of infection between flats in high-rise residential buildings: Particle simulation, Building and Environment 44 (2) (2009) 402–410.

[11] C. He, L. Morawska, D. Gilbert, Particle deposition rates in residential houses, Atmospheric Environment 39 (21) (2005) 3891–3899.

[12] W. J. Riley, T. E. McKone, A. C. Lai, W. W. Nazaroff, Indoor particulate matter of outdoor origin: importance of size-dependent removal mechanisms, Environmental science & technology 36 (2) (2002) 200–207.

[13] J. G. Crump, J. H. Seinfeld, Turbulent deposition and gravitational sedimentation of an aerosol in a vessel of arbitrary shape, Journal of Aerosol Science 12 (5) (1981) 405–415.

[14] S. L. Miller, W. W. Nazaroff, J. L. Jimenez, A. Boerstra, G. Buonanno, S. J. Dancer, J. Kurnitski, L. C. Marr, L. Morawska, C. Noakes, Transmission of SARS-CoV-2 by inhalation of respiratory aerosol in the Skagit Valley Chorale superspreading event, Indoor Air 00 (2020) 1–10. arXiv:https://onlinelibrary.wiley.com/doi/pdf/10.1111/ina.12751, doi:10.1111/ina.12751.

[15] E. C. Riley, G. Murphy, R. L. Riley, AIRBORNE SPREAD OF MEASLES IN A SUBURBAN ELEMENTARY SCHOOL, American Journal of Epidemiology 107 (5) (1978) 421–432. arXiv:https://academic.oup.com/aje/article-pdf/107/5/421/126698/107-5-421.pdf, doi:10.1093/oxfordjournals.aje.a112560.

[16] W. W. Nazaroff, M. Nicas, S. L. Miller, Framework for Evaluating Measures to Control Nosocomial Tuberculosis Transmission, Indoor Air 8 (4) (1998) 205–218. arXiv:https://onlinelibrary.wiley.com/doi/pdf/10.1111/j.1600-0668.1998.00002.x, doi:10.1111/j.1600-0668.1998.00002.x.

[17] K. P. Fennelly, E. A. Nardell, The Relative Efficacy of Respirators and Room Ventilation in Preventing Occupational Tuberculosis, Infection Control & Hospital Epidemiology 19 (10) (1998) 754–759. doi:10.2307/30141420.

[18] M. Nicas, W. W. Nazaroff, A. Hubbard, Toward Understanding the Risk of Secondary Airborne Infection: Emission of Respirable Pathogens, Journal of Occupational and Environmental Hygiene 2 (2005) 143–154. doi:10.1080/15459620590918466.

[19] G. Buonanno, L. Stabile, L. Morawska, Estimation of airborne viral emission: Quanta emission rate of SARS-CoV-2 for infection risk assessment, Environment International 141 (2020) 105794. doi:https://doi.org/10.1016/j.envint.2020.105794. URL https://www.sciencedirect.com/science/article/pii/S0160412020312800

[20] A. C. Lai, W. W. Nazaroff, Modeling indoor particle deposition from turbulent flow onto smooth surfaces, Journal of aerosol science 31 (4) (2000) 463–476.

[21] C. He, L. Morawska, D. Gilbert, Particle deposition rates in residential houses, Atmospheric Environment 39 (21) (2005) 3891–3899.

[22] M. Nicas, Estimating exposure intensity in an imperfectly mixed room, American Industrial Hygiene Association Journal 57 (6) (1996) 542–550.

[23] J. W. Cherrie, The effect of room size and general ventilation on the relationship between near and far-field concentrations, Applied occupational and environmental hygiene 14 (8) (1999) 539–546.

[24] J. W. Cherrie, L. Maccalman, W. Fransman, E. Tielemans, M. Tischer, M. Van Tongeren, Revisiting the effect of room size and general ventilation on the relationship between near-and far-field air concentrations, Annals of occupational hygiene 55 (9) (2011) 1006–1015.

[25] M. Nicas, J. Neuhaus, Predicting benzene vapor concentrations with a near field/far field model, Journal of occupational and environmental hygiene 5 (9) (2008) 599–608.

[26] S. F. Arnold, Y. Shao, G. Ramachandran, Evaluating well-mixed room and near-field–far-field model performance under highly controlled conditions, Journal of Occupational and Environmental Hygiene 14 (6) (2017) 427–437.

[27] S. F. Arnold, Y. Shao, G. Ramachandran, Evaluation of the well mixed room and near-field far-field models in occupational settings, Journal of occupational and environmental hygiene 14 (9) (2017) 694–702.

[28] H. Sajjadi, M. Salmanzadeh, G. Ahmadi, S. Jafari, Simulations of indoor airflow and particle dispersion and deposition by the lattice Boltzmann method using LES and RANS approaches, Building and Environment 102 (2016) 1–12.

[29] N. Gao, J. Niu, L. Morawska, Distribution of respiratory droplets in enclosed environments under different air distribution methods, Building Simulation 1 (2008) 326–335.

[30] B. Zhao, C. Chen, Z. Tan, Modeling of ultrafine particle dispersion in indoor environments with an improved drift flux model, Journal of Aerosol Science 40 (1) (2009) 29–43.

[31] D. Mage, W. Ott, Accounting for Nonuniform Mixing and Human Exposure in Indoor Environments, In STP1287-EB Characterizing Sources of Indoor Air Pollution and Related Sink Effects, ed. B. Tichenor (1996).263–278.

[32] N. E. Klepeis, Validity of the Uniform Mixing Assumption: Determining Human Exposure to Environmental Tobacco Smoke, Environmental Health Perspectives 107 (1999) 357–363.

[33] D. Licina, Y. Tian, W. W. Nazaroff, Inhalation intake fraction of particulate matter from localized indoor emissions, Building and Environment 123 (2017) 14–22. doi:https://doi.org/10.1016/j.buildenv.2017.06.037.

[34] J. Curtius, M. Granzin, J. Schrod, Testing mobile air purifiers in a school classroom: Reducing the airborne transmission risk for SARS-CoV-2, Aerosol Science and Technology 55 (5) (2021) 586–599.

[35] E. S. Mousavi, N. Kananizadeh, R. A. Martinello, J. D. Sherman, COVID-19 Outbreak and Hospital Air Quality: A Systematic Review of Evidence on Air Filtration and Recirculation, Environmental Science & Technology 55 (7) (2021) 4134–4147.

[36] Y.-F. Ren, Q. Huang, T. Marzouk, R. Richard, K. Pembroke, P. Martone, T. Venner, H. Malmstrom, E. Eliav, Effects of mechanical ventilation and portable air cleaner on aerosol removal from dental treatment rooms, Journal of Dentistry 105 (2021) 103576.

[37] M. Alavy, J. A. Siegel, In-situ effectiveness of residential HVAC filters, Indoor Air 30 (1) (2020) 156–166.

[38] W. Ott, N. Klepeis, P. Switzer, Air change rates of motor vehicles and in-vehicle pollutant concentrations from secondhand smoke, Journal of Exposure Science & Environmental Epidemiology 18 (3) (2008) 312–325.

[39] T. Hussein, T. Glytsos, J. Ondráček, P. Dohányosová, V. Ždímal, K. Hämeri, M. Lazaridis, J. Smolík, M. Kulmala, Particle size characterization and emission rates during indoor activities in a house, Atmospheric Environment 40 (23) (2006) 4285–4307. doi:https://doi.org/10.1016/j.atmosenv.2006.03.053.

[40] Y. Ishizu, General equation for the estimation of indoor pollution, Environmental Science & Technology 14 (10) (1980) 1254–1257.

[41] D. Licina, J. Pantelic, A. Melikov, C. Sekhar, K. W. Tham, Experimental investigation of the human convective boundary layer in a quiescent indoor environment, Building and Environment 75 (2014) 79–91.

[42] D. Rim, A. Novoselac, Transport of particulate and gaseous pollutants in the vicinity of a human body, Building and Environment 44 (9) (2009) 1840–1849.

[43] Z. Cheng, A. Aganovic, G. Cao, Z. Bu, Experimental and simulated evaluations of airborne contaminant exposure in a room with a modified localized laminar airflow system, Environmental Science and Pollution Research (2021).

[44] J. Qian, A. R. Ferro, K. R. Fowler, Estimating the Resuspension Rate and Residence Time of Indoor Particles, Journal of the Air & Waste Management Association 58 (4) (2012) 502–516.

[45] A. C. Lai, K. Wang, F. Chen, Experimental and numerical study on particle distribution in a two-zone chamber, Atmospheric Environment 42 (8) (2008) 1717–1726.

[46] W. R. Ott, N. E. Klepeis, P. Switzer, Analytical Solutions to Compartmental Indoor Air Quality Models with Application to Environmental Tobacco Smoke Concentrations Measured in a House, Journal of the Air & Waste Management Association 53 (8) (2003) 918–936.

[47] S. Miller, W. Nazaroff, Environmental tobacco smoke particles in multizone indoor environments, Atmospheric Environment 35 (12) (2001) 2053–2067. doi:https://doi.org/10.1016/S1352-2310(00)00506-9.

[48] C. Poon, L. Wallace, A. C. Lai, Experimental study of exposure to cooking emitted particles under single zone and two-zone environments, Building and Environment 104 (2016) 122–130. doi:https://doi.org/10.1016/j.buildenv.2016.04.026.

[49] G. Bagheri, E. Bodenschatz, B. Hejazi, O. Schlenczek, B. Thiede, Infection risk in large indoor environments, Manuscript submitted and available upon request (2021).

[50] S. M. Kissler, J. R. Fauver, C. Mack, C. Tai, M. Breban, A. E. Watkins, R. Samant, D. Anderson, D. Ho, N. D. Grubaugh, et al., Densely sampled viral trajectories suggest longer duration of acute infection with B. 1.1. 7 variant relative to non-B. 1.1. 7 SARS-CoV-2, medRxiv (2021).

[51] HEADS — Human Emission of Aerosol and Droplet Statistics, https://aerosol.ds.mpg.de, accessed: 2021-05-04.

[52] International Commission on Radiological Protection (Ed.), Human respiratory tract model for radiology protection: a report of a Task Group of the International Commission on Radiological Protection, 1st Edition, no. 66 in ICRP publication, Pergamon, Oxford, 1994, oCLC: 832441728.

